# CONORM: Context-Aware Entity Normalization for Adverse Drug Event Detection

**DOI:** 10.1101/2023.09.26.23296150

**Authors:** Anthony Yazdani, Hossein Rouhizadeh, Alban Bornet, Douglas Teodoro

**Affiliations:** Department of Radiology and Medical Informatics, University of Geneva, Geneva, Switzerland

**Keywords:** Adverse Drug Events, Named Entity Recognition, Entity Normalization, Entity Resolution, Large Language Models

## Abstract

Adverse drug events (ADEs) are a critical aspect of patient safety and pharmacovigilance, with significant implications for patient outcomes and public health monitoring. The increasing availability of electronic health records, social media, and online patient forums provides valuable yet challenging unstructured data sources for ADE surveillance. To address these challenges, we introduce CONORM, a novel framework integrating named entity recognition (NER) and entity normalization (EN) for ADE resolution across diverse textual domains. CONORM comprises CONORM-NER and CONORM-EN, featuring a dual-encoder architecture with dynamic context refining (DCR). The DCR mechanism adaptively combines isolated entity embeddings with contextual representations. Our analyses demonstrate this approach effectively adjusts model behavior according to text formality, enhances precision on out-of-distribution concepts, and substantially reduces normalization errors compared to context-agnostic baselines. CONORM was evaluated on tweets, forum posts, and structured product labels, achieving end-to-end F1-scores of 63.86%, 72.45%, and 84.99%, respectively, surpassing existing solutions by an average margin of 35%. These results highlight CONORM’s robust adaptability across domains, enabled by DCR’s effective context utilization. CONORM offers a scalable, reproducible solution for pharmacovigilance, with pre-computed target embeddings enhancing inference efficiency. Its generalization establishes it as a robust tool for ADE surveillance. Source code is publicly available at https://github.com/ds4dh/CONORM.

## Introduction

Adverse drug events (ADEs) are unintended injuries resulting from medication use, whether appropriate or inappropriate, and they represent a substantial portion of adverse events in healthcare ^7,2^. ADEs pose a significant public health issue, leading to considerable morbidity and hospitalization ^3^. They are among the leading causes of hospital-related complications in developed countries, and their impact is expected to increase due to aging populations and rising medication use ^4–6^. The economic burden of ADEs is substantial, since it involves increased healthcare utilization, prolonged hospital stays, and lost productivity ^7^. For example, in the United Kingdom, ADEs cost the national health service approximately £99 million per year and account for over 181,000 bed-days and more than 1,700 deaths annually ^8^. In the United States, estimates from the 1990s placed the costs of serious ADEs between $30 billion and $137 billion annually ^9^, amounts that have more than doubled since then ^10^. These statistics highlight the need for effective pharmacovigilance strategies to identify ADEs to support drug-related morbidity and mortality prevention.

Named entity recognition (NER) and entity normalization (EN) are important natural language processing techniques in pharmacovigilance and play a significant role in ADE information extraction. NER involves identifying entities such as drug names, symptoms, and conditions within unstructured text, which is necessary for extracting potential ADE mentions from vast data sources like electronic health records (EHRs) and social media platforms ^11–13^. EN focuses on standardizing these identified entities to controlled vocabulary terms, such as those in medical ontologies, like the Medical Dictionary for Regulatory Activities (MedDRA) (https://www.meddra.org/). This normalization ensures that diverse expressions of the same ADE are uniformly represented to facilitate accurate aggregation and analysis, as well as semantic interoperability.

Traditional methods for documenting ADEs are limited by under-reporting ^14,15^. In contrast, a more comprehensive understanding of ADEs can be achieved by considering diverse data sources, which range from highly informal social media posts to formal clinical narratives ^11–13,16,17^. Social media provide real-time, patient-generated insights into ADEs that might otherwise go unreported, while clinical narratives in EHRs offer detailed information from healthcare professionals. However, both types of sources present challenges for ADE detection due to their unstructured nature and variability in language. Social media data is often marked by slang, abbreviations, misspellings, and colloquial expressions^18^, whereas clinical narratives rely on specialized medical terminology ^19–21^. Despite the progress made in text normalization techniques, the dynamic language of social media and the complexity of medical jargon in clinical texts still require continuous refinement. Hence, there is a need for advanced methods capable of generalizing and accurately extracting and normalizing ADE mentions from unstructured text across varying levels of formality.

In response to these challenges, we introduce CONORM, a context-aware entity normalization framework that provides an end-to-end solution for ADE detection and normalization across various text domains. CONORM integrates a fine-tuned discriminative language model for ADE recognition (CONORM-NER) with dynamic context refining (DCR) in its normalization module (CONORM-EN). CONORM-EN leverages the DCR mechanism to enhance entity normalization by dynamically balancing isolated entity embeddings (capturing general-domain, pre-trained semantics) and contextual embeddings (capturing contextual, task-specific information). This DCR process is empirically shown to adapt near-optimally to text formality (Section), enhance generalization by improving precision on out-of-distribution concepts (Section, Section), and act as an effective error correction module over baseline context-free approaches (Section). Additionally, we constructed CONORM-EN to employ embedding pre-computation for scalable inference over large terminologies like MedDRA (Section), making the system inherently compatible with vector databases from pre-training to inference. This overall approach allows CONORM to effectively handle the variability and complexity of language in unstructured text sources, including highly informal social media posts, semi-formal patient forums, and formal clinical narratives. Our analyses confirm this adaptability across varying text formalities, as well as different context and mention lengths (Section, Section).

While generative large language models (LLMs) have demonstrated broad capabilities, their application to specialized tasks like fine-grained biomedical NER and EN presents challenges. Studies indicate that smaller, specialized models achieve superior NER performance compared to generative LLMs like ChatGPT and UniversalNER ^22,23^. Furthermore, recent studies highlight how generative LLMs struggle to encode medical concepts for biomedical concept normalization tasks ^24^. This suggests that the significant computational cost of generative LLMs does not always translate to optimal performance for specific, structured information extraction tasks. CONORM is designed to bridge this gap by leveraging efficient encoder-based transformer architectures. Its DCR mechanism, leveraging attention and embedding fusion principles ^25,26^, yields a notable efficiency advantage. Because the isolated entity encoder is frozen, target concept embeddings can be pre-computed before main training and remain valid for inference. This offers enhanced efficiency compared to generative LLMs and bi-encoder systems ^27^, in which pre-computation is performed only post-training.

The main contributions of this work are:

- A novel end-to-end framework, CONORM, for ADE detection and normalization across diverse text domains.
- The dynamic context refining (DCR) mechanism, which adaptively integrates isolated and contextual entity representations, enhancing normalization accuracy and generalization.
- State-of-the-art end-to-end performance achieved on three benchmark datasets representing informal, semi-formal, and formal text.
- A comprehensive analysis characterizing CONORM’s performance, including its adaptive context balancing, robustness to context and mention length variations, generalization to unseen concepts, and impact on error correction.
- A publicly available implementation promoting reproducibility (https://github.com/ds4dh/CONORM).

By addressing these challenges, CONORM enhances pharmacovigilance efforts, enabling more accurate detection and normalization of ADE mentions in unstructured texts. Our results and analyses demonstrate that CONORM advances the state-of-the-art in ADE detection and normalization, offering a robust, efficient, and generalizable framework for improving drug safety monitoring.

### Related work

Early tools in biomedical text processing, such as MetaMap ^28^, were developed to map biomedical text to concepts in the Unified Medical Language System (UMLS). MetaMap uses linguistic and symbolic processing to identify medical concepts from text. While it has been widely used for biomedical text mining, MetaMap faces significant limitations when dealing with informal language, slang, and large datasets due to its reliance on lexical lookup and rule-based methods. Its performance decreases in unstructured or colloquial text, such as social media posts, and it struggles with scalability issues when processing large volumes of data^29,30^. To overcome some of MetaMap’s limitations, researchers developed tools like MedCAT^31^, an open-source medical concept annotation toolkit. MedCAT leverages unsupervised machine learning and ontologies like SNOMED-CT and UMLS to annotate medical concepts in unstructured text. It has shown improved performance in extracting concepts from EHRs and supports customization through an annotation interface.

The advent of transformer architectures advanced natural language processing, with models like BERT^32^ introducing attention-based contextual embeddings that capture rich bidirectional language patterns. In the biomedical domain, BioBERT^33^ extended this approach by pre-training on large-scale biomedical corpora to enhance the model’s understanding of complex biomedical language. These models have demonstrated promising results in ADE extraction tasks ^34^, enabling more accurate identification of medical entities in unstructured text. Researchers have also effectively applied transformer-based models to social media texts. DeepADEMiner ^35^, for instance, is a multi-step pipeline based on RoBERTa ^36^ for filtering ADE-related tweets and on BERT-based models for span extraction and normalization. This approach has shown success in handling informal language and extracting ADEs from social media data. Hybrid approaches have also been explored, integrating models like *mcn-en-smm4h* with DeepADEMiner’s span extraction to improve normalization of ADE mentions to MedDRA terms ^37^. Additionally, the use of text-to-text transformer models like T5^38^ has been investigated for tweet-based ADE extraction by framing the task as a sequence-to-sequence problem. Kanagasabai *et al*. ^39^ complemented this approach with cosine similarity-based retrieval using sentence transformers ^40^ for normalizing ADE spans. Together, these studies highlight the versatility of transformer architectures and pre-trained language models in both extraction and normalization tasks.

Scalability is another critical concern in ADE normalization efforts. To address this issue, Ul Haq *et al*. ^41^ developed a scalable, production-ready NLP pipeline capable of processing ADEs from vast amounts of unstructured text, including social media data. Their approach combines a state-of-the-art NER model for ADE extraction with two relation extraction models, one of which is based on BioBERT. Their pipeline, implemented in Apache Spark ^42^, emphasizes scalability and enables the processing of large datasets. To improve computational efficiency, the JNRF model ^43^ was proposed as an end-to-end solution for ADE NER and relation extraction in variable-length documents. JNRF is based on weight-shared Fourier networks for low-complexity token mixing, which reduces computational costs and memory consumption. While it demonstrates faster training times and reduced resource usage, JNRF primarily addresses clinical text and may face limitations when applied to informal or highly variable text sources like social media.

Traditional NER systems are often restricted to predefined entity types, limiting their flexibility and pre-training potential. GLiNER ^23^ addresses this limitation by using a bidirectional transformer encoder to extract arbitrary entity types. By matching entity embeddings with textual spans in a shared latent space, GLiNER enables open NER, outperforming large language models like ChatGPT in zero-shot evaluations^23^.

However, GLiNER’s joint encoding of text and entities can lead to computational overhead, particularly when dealing with large ontologies like MedDRA. To overcome these limitations, GLiNER-bi ^27^ introduces a bi-encoder architecture that decouples text and entity encodings into separate transformers. This approach enables the pre-computation of entity embeddings at inference time, reducing computational overhead and improving efficiency.

To further address the complexities of ADE normalization in informal texts, Remy *et al*. ^44^ proposed using a general-purpose model initialization via BioLORD and to fine-tune the model with semantic text similarity. This approach leverages both general-domain and domain-specific pre-training to improve ADE normalization in social media contexts. Their approach outperformed models pre-trained exclusively on biomedical corpora. The success of BioLORD-STS, particularly in zero-shot scenarios, highlights the potential of combining diverse pre-training strategies to handle variability and informality in language. Similarly, in our previous work ^45^, we presented a two-stage pipeline for ADE resolution that ranked first in the Social Media Mining for Health Research and Applications (SMM4H) 2023 shared task 5^13^. First, we fine-tuned BERTweet ^46^ for NER to extract ADE mentions from tweets. In a second stage, we combine similarity rankings from multiple general-domain and specialized sentence transformers using a zero-shot normalization approach based on reciprocal-rank fusion ^47,48^.

## Materials and methods

### Datasets

In this study, we used three benchmark datasets to evaluate the effectiveness of CONORM, focusing specifically on continuous ADE entities. These datasets were chosen to cover a wide spectrum of text formality and complexity, ensuring a robust assessment of our solution. The datasets included are the Social Media Mining for Health (SMM4H) 2023 shared task 5^13^, the CSIRO Adverse Drug Event Corpus (CADEC) ^16^, and the Text Analysis Conference (TAC) 2017 dataset ^17^, each presenting unique challenges and use cases for our model. Evaluation for each dataset is conducted at the preferred term (PT) level, which represents the most granular level of distinct concepts within the MedDRA ontology. System organ classes (SOCs) provide the highest-level groupings of these PTs. Together, these datasets enable a comprehensive evaluation of ADE detection and normalization across diverse linguistic, structural, and domain-specific contexts.

#### SMM4H 2023

The main challenge of the SMM4H 2023 dataset is its informal nature, including slang, misspellings and abbreviations. This dataset is widely used for the normalization of ADEs in English tweets and serves as a standard benchmark for resolving ADE mentions to their corresponding MedDRA terms. It comprises approximately 18,000 labeled tweets for training and 10,000 for testing. The evaluation metrics assess system performance across all MedDRA PTs and the model’s generalization to out-of-distribution ADEs. To ensure robust model evaluation, the test set labels remain undisclosed. Participants submit their predictions through CodaLab (https://codalab.org/) for evaluation. For our experiments, we split the official training data into separate training (90%) and validation (10%) sets, using the official validation set as our internal test set. We used the official test tweets to evaluate our model on the undisclosed labels via CodaLab. Detailed statistics for the SMM4H 2023 dataset are presented in Table 1. The average tweet length across the splits is approximately 16 words, with tweets typically comprising around two sentences. However, the low entity-to-sentence ratio (e.g., 0.06 in the training set) highlights the sparsity of ADE mentions, posing challenges for entity resolution within noisy and unrelated content. Moreover, the dataset’s SOC distribution reveals significant skewness, with a high prevalence of *Psychiatric, General*, and *Nervous system* disorders. In contrast, SOCs such as *Reproductive* and *Cardiac* disorders are scarcely represented among those present in all data splits.

**Table 1.** Summary statistics of the SMM4H 2023 dataset, including descriptive metrics and the distribution of entities across the top 5 and least 5 system organ classes (SOCs) in the training, validation, and test sets. Only non-zero SOCs in all splits are shown.

| Metric | Train | Validation | Test |
| --- | --- | --- | --- |
| Total entities | 1570 | 138 | 87 |
| Average word count | 16 | 16 | 16 |
| Average sentence count | 2 | 2 | 2 |
| Average entity-to-sentence ratio | 0.06 | 0.05 | 0.06 |
| **Entities per SOC** |  |  |  |
| **Top 5 SOCs** |  |  |  |
| Psychiatric disorders | 667 | 66 | 39 |
| General disorders and administration site conditions | 488 | 47 | 20 |
| Nervous system disorders | 466 | 40 | 31 |
| Musculoskeletal and connective tissue disorders | 103 | 10 | 3 |
| Gastrointestinal disorders | 88 | 8 | 8 |
| **Least 5 SOCs** |  |  |  |
| Reproductive system and breast disorders | 16 | 2 | 1 |
| Cardiac disorders | 29 | 1 | 1 |
| Vascular disorders | 35 | 3 | 2 |
| Injury, poisoning and procedural complications | 40 | 3 | 2 |
| Skin and subcutaneous tissue disorders | 67 | 2 | 6 |

#### CADEC

The CADEC dataset comprises annotated medical forum posts detailing patient-reported ADEs. These posts, written in colloquial language, often deviate from formal grammar and punctuation. Annotations link ADE mentions to MedDRA PTs. The dataset consists of 1,250 files, which we split into 875 files for training, 187 for validation, and 188 for testing. CADEC’s semi-formal text provides a middle ground between the highly informal SMM4H 2023 and the formal TAC datasets, which allows testing our models in a varied linguistic environment. Detailed statistics for CADEC are shown in Table 2. The average word count of posts, around 80-90 words across splits, is significantly higher than that of SMM4H 2023, offering richer context for ADE resolution, although the average sentence count remains relatively low. The average entity-to-sentence ratio (e.g., 0.64 in the training set) reflects a moderately dense annotation scheme. The SOC distribution highlights a strong dominance of *Musculoskeletal, Nervous system*, and *General* disorders. In contrast, SOCs such as *Endocrine* and *Social circumstances* disorders are scarcely represented among those present in all data splits.

**Table 2.** Summary statistics of the CADEC dataset, showing descriptive metrics and the distribution of entities across the top 5 and least 5 system organ classes (SOCs) in the training, validation, and test sets. Only non-zero SOCs in all splits are shown.

| Metric | Train | Validation | Test |
| --- | --- | --- | --- |
| Total entities | 4087 | 718 | 834 |
| Average word count | 86 | 74 | 79 |
| Average sentence count | 7 | 7 | 7 |
| Average entity-to-sentence ratio | 0.64 | 0.58 | 0.66 |
| **Entities per SOC** |  |  |  |
| **Top 5 SOCs** |  |  |  |
| Musculoskeletal and connective tissue disorders | 1101 | 191 | 168 |
| Nervous system disorders | 1000 | 159 | 193 |
| General disorders and administration site conditions | 885 | 175 | 181 |
| Psychiatric disorders | 616 | 107 | 125 |
| Gastrointestinal disorders | 443 | 85 | 95 |
| **Least 5 SOCs** |  |  |  |
| Endocrine disorders | 2 | 1 | 4 |
| Social circumstances | 5 | 4 | 2 |
| Hepatobiliary disorders | 13 | 2 | 5 |
| Immune system disorders | 15 | 3 | 7 |
| Ear and labyrinth disorders | 41 | 7 | 5 |

#### TAC

The TAC dataset is designed to evaluate information extraction performance from structured product labels (SPLs). In contrast with the informal language of social media posts, this dataset is valuable for assessing the model’s performance on medical texts. The training set includes 101 SPLs, and the test set 99 SPLs. We used 90 of the official training SPLs for training and 11 for validation. The structured and formal nature of the TAC dataset provides a stringent test for our model’s capability to handle formal medical terminologies and well-structured documents. As shown in Table 3, the average length of SPLs is significantly longer than the other datasets, with average word counts around 900-1000 per sample across the splits. This extensive length offers rich contextual information. The dense nature of these texts is reflected in the high entity-to-sentence ratio (e.g., 1.50 in the training set) and the sheer number of annotations, underscoring the need for the model to handle frequent ADE mentions within complex documents. The SOC distribution is notably more balanced compared to SMM4H 2023 and CADEC, with categories such as *Nervous system, General*, and *Gastrointestinal* disorders predominating, ensuring a comprehensive evaluation across diverse clinical contexts. While this balance is a strength for robust evaluation, some categories like *Social circumstances* and *Ear and labyrinth* disorders remain less frequent.

**Table 3.** Summary statistics of the TAC dataset, presenting descriptive metrics and the distribution of entities across the top 5 and least 5 system organ classes (SOCs) in the training, validation, and test sets. Only non-zero SOCs in all splits are shown.

| Metric | Train | Validation | Test |
| --- | --- | --- | --- |
| Total entities | 10630 | 1711 | 11174 |
| Average word count | 892 | 1060 | 919 |
| Average sentence count | 34 | 41 | 35 |
| Average entity-to-sentence ratio | 1.50 | 1.62 | 1.36 |
| **Entities per SOC** |  |  |  |
| **Top 5 SOCs** |  |  |  |
| Nervous system disorders | 1538 | 163 | 1609 |
| General disorders and administration site conditions | 1401 | 219 | 1526 |
| Gastrointestinal disorders | 1287 | 255 | 1477 |
| Vascular disorders | 1265 | 116 | 1271 |
| Infections and infestations | 1057 | 225 | 820 |
| **Least 5 SOCs** |  |  |  |
| Social circumstances | 4 | 2 | 4 |
| Ear and labyrinth disorders | 45 | 1 | 40 |
| Pregnancy, puerperium and perinatal conditions | 57 | 11 | 89 |
| Hepatobiliary disorders | 191 | 18 | 292 |
| Neoplasms benign, malignant and unspecified | 208 | 81 | 251 |

### CONORM-NER: named entity recognition

NER is a key component of end-to-end entity resolution. In our study, CONORM-NER is responsible for identifying ADEs from text data, which is necessary for accurate normalization, especially when multiple ADE mentions are present in a text. An overview of CONORM-NER is shown in Figure 1A.

**Figure 1.**
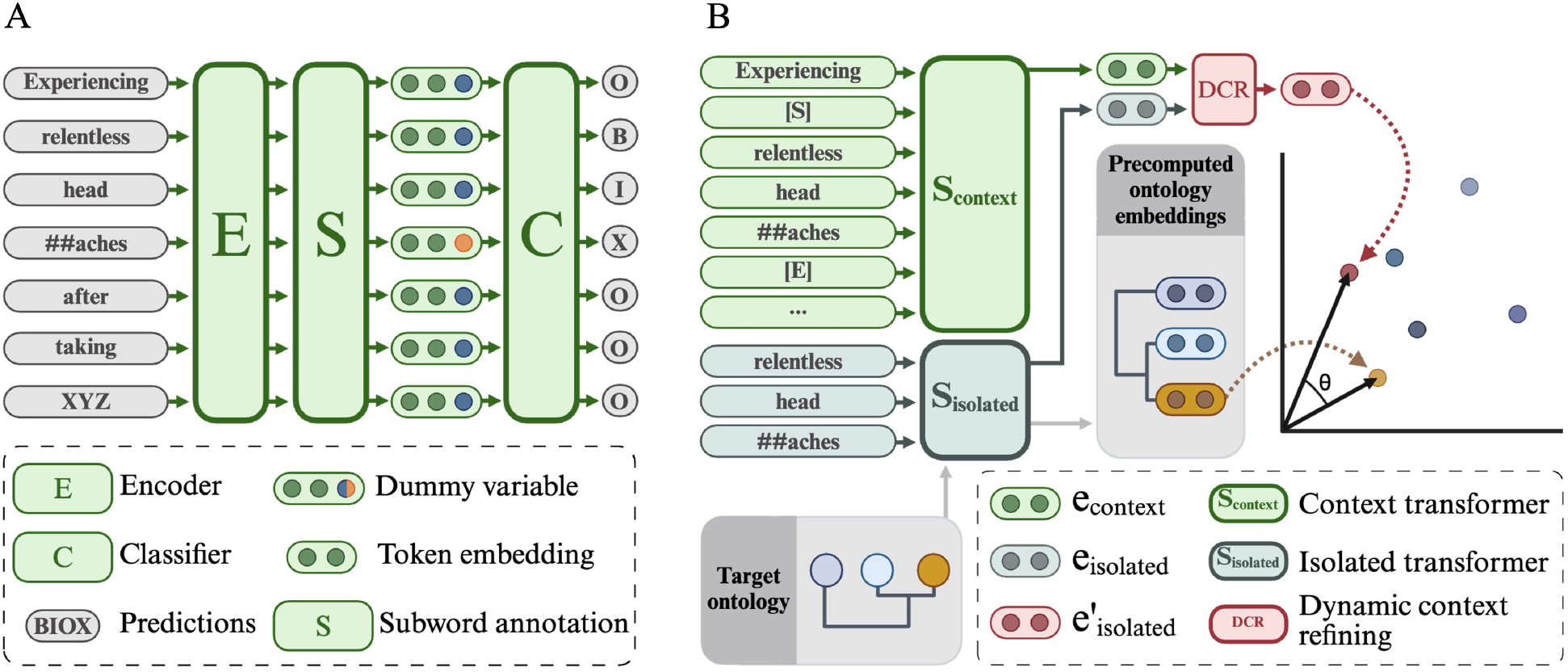
Schematic overview of the CONORM framework: (A) CONORM-NER for named entity recognition and (B) CONORM-EN for entity normalization, illustrating the integration of dynamic context refining (DCR) and dual-encoder architectures.

We start by splitting raw texts into words and punctuation symbols using regular expressions. These pre-tokenized texts are then processed through the tokenizer of the transformer encoder backbone. We adopt this two-stage tokenization strategy to ensure that our labeling method aligns closely with the annotated dataset. Each text passage is labeled using the BIOX scheme, “B” for the beginning of an ADE mention, “I” for the inside, “O” for the outside, and “X” for continuation sub-words, regardless of their relation to an ADE ^45^.

Given an input sequence *x*, CONORM-NER generates a sequence of hidden states:

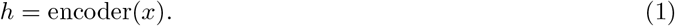

To mitigate overfitting, we apply a 10% dropout layer to these hidden states:

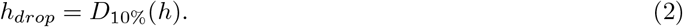

Sub-words pose a unique challenge during classification. When a word is segmented into multiple sub-words, we distinguish the leading sub-word from the continuation sub-words. Leading sub-words are labeled as “B”, “I”, or “O”, while all continuation sub-words are labeled as “X”. The labels of continuation sub-words are deterministic based on the tokenization process. However, these sub-words must be retained during the forward pass for proper contextualization of the input sequence.

To prevent the model from learning to identify continuation sub-words from deep features (a deterministic task), we introduce a dummy variable *s*. This variable is set to 1 for continuation sub-words and 0 for all other token types ^45^. Each token’s contextual embedding is then concatenated with its respective dummy variable before being fed into the classification layer:

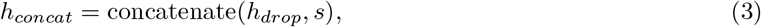

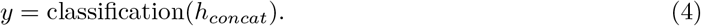

CONORM-NER employs a two-phase learning strategy. During the first epoch, only the classification layer weights are trained, leaving the pre-trained parameters unchanged. This allows the model to refine the randomly initialized layers before back-propagating errors to the pre-trained weights. From the second epoch onwards, all parameters are available for gradient updates.

### CONORM-EN: entity normalization

The challenge of extracting ADEs from textual data goes beyond entity recognition. It is also crucial to map recognized entities to standardized terminologies in medical databases for a unified understanding and further analysis. CONORM-EN, shown in Figure 1B, is our dedicated sub-model for this entity normalization task.

In CONORM-EN, the context is defined as the sentence containing the target entity to normalize. However, for sufficiently short documents that fit within the model’s input size, the entire document may be used as context. Our approach uses two transformer tokenizers, one for context and another for isolated entities. This two-tokenizer strategy is integral to CONORM-EN’s architecture. One sentence transformer interprets the broader context surrounding the identified entity, while the second focuses solely on the entity itself. We introduce special tokens, [*S*] for start and [*E*] for end, to mark the boundaries of target entities within their context. These tokens highlight the target entity, especially in the presence of non-target entities in the context. They are integrated into the context tokenizer’s vocabulary and are assigned unique embeddings in the context sentence transformer’s embedding matrix.

Consider a text *t* describing an ADE in context, such as “Experiencing *relentless headaches* after taking XYZ”. Using CONORM-NER, an ADE passage *p*_*ade*_ is extracted and identified from *t*, i.e., “relentless headaches”. Then, the text *t* is transformed for CONORM-EN to encapsulate the ADE passage with special tokens, creating a new tagged text *c*_*ade*_, i.e., “Experiencing [S] relentless headaches [E] after taking XYZ”. The context tokenizer processes the entire tagged text *c*_*ade*_, while the isolated entity tokenizer only processes the ADE passage *p*_*ade*_:

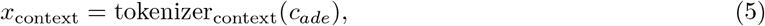

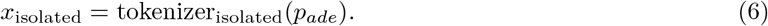

### Dynamic context refining

CONORM-EN constructs a representation of the target entity using dynamic context refining (DCR). Contextual and isolated entity embeddings are integrated in a late fusion stage supported by a simplified attention mechanism with constant *𝒪* (*d*) time and space complexity, where *d* is the embedding dimensionality. Given a token sequence from a context *c*_*ade*_, i.e., *x*_context_ and a token sequence from an isolated entity *p*_*ade*_, i.e., *x*_isolated_, we compute their respective embedding vectors by averaging the hidden states from two sentence transformers, *S*_context_ and *S*_isolated_, as follows:

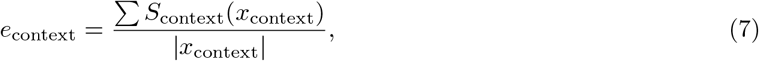

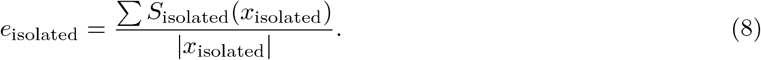

DCR then refines *e*_isolated_ by incorporating insights from *e*_context_, as follows:

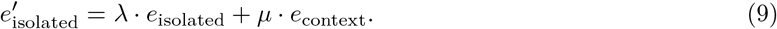

Here, *λ* and *µ* are weights derived using a softmax function:

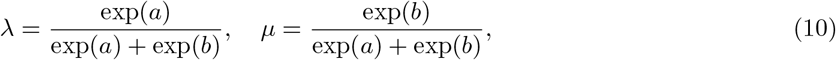

where *λ >* 0, *µ >* 0, and *λ* + *µ* = 1. The parameters *a* and *b* that influence *λ* and *µ* are defined as:

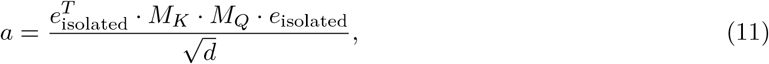

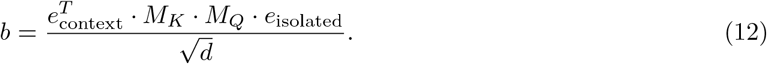

In CONORM-EN, the context sentence transformer *S*_context_ and the *M* matrices (i.e., *M*_*K*_ and *M*_*Q*_) are trainable parameters, allowing the model to learn and adapt to task-specific patterns. Conversely, the isolated entity sentence transformer *S*_isolated_ remains frozen with deactivated dropout layers, ensuring consistent operation in inference mode, even during model training. The coefficients *λ* and *µ* act as dynamic weighting factors, determining the influence of the respective embeddings. Specifically, *λ* represents the model’s reliance on the isolated entity embedding, capturing static semantic knowledge from the pre-trained transformer, while *µ* dictates the contribution of the contextual embedding, incorporating task-specific information. The learnable matrices *M*_*K*_ and *M*_*Q*_ facilitate these interactions through a simplified attention mechanism, using the embeddings as values. Together, this architecture enables CONORM-EN to dynamically balance static general knowledge and task-specific contextual information.

### Scalable inference

CONORM-EN achieves efficient ADE normalization by pre-computing target embeddings and decoupling the forward pass from the target vocabulary. We pre-compute embeddings for the target vocabulary using *S*_isolated_, which eliminates the need for runtime recomputation and significantly reduces computational costs. During inference, CONORM-EN’s forward pass focuses solely on input text, operating independently of the vocabulary. This decoupled architecture supports integration with vector databases, enabling efficient retrieval through techniques such as locality-sensitive hashing, random projection, and quantization.

The inference process begins with embedding each ADE mention, i.e., 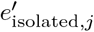, identified by CONORM-NER. These embeddings are compared to the pre-computed target embeddings 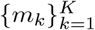, where *K* is the total number of concepts in the vocabulary. The cosine similarity is then used to quantify the alignment between an ADE mention and each target embedding:

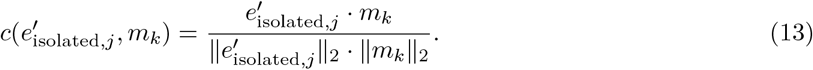

Here,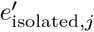 is the embedding of the *j*-th ADE mention, and *m*_*k*_ is the embedding of the *k*-th target concept. For each mention, we compute a similarity score vector:

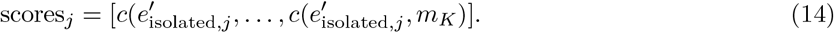

The concept corresponding to the highest similarity score in this vector is selected as the predicted normalization for the ADE mention.

### Robust training

To train CONORM-EN, we generate dissimilar pairs through balanced negative sampling. In this context, a positive pair refers to a combination 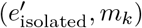 where *m*_*k*_ is the correct concept embedding for the target entity passage *p*_*ade*_. The number of positive pairs is dictated by the annotations available in the training set, which specify the correct mappings for each ADE. Conversely, a negative pair consists of 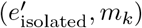 where *m*_*k*_ is not the gold label for *p*_*ade*_. Negative pairs are created using negative sampling, which involves randomly selecting a label from the set of all possible labels that are not explicitly marked as correct. The quality of these negative pairs is uncertain because the sampling process is uniform, meaning that we might sample a label that is semantically similar to the true label. For example, in MedDRA, “Headache” might be closely related to “Migraine,” leading to ambiguity in the negative sampling process.

To address this challenge, we use the cosine embedding loss *L*, which is less sensitive to noisy negative examples:

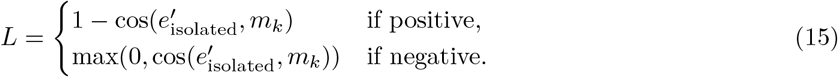

For positive pairs, the loss function ensures that the cosine similarity between 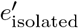 and its corresponding concept embedding *m*_*k*_ approaches 1.0. For negative pairs, the loss function only contributes if the cosine similarity is greater than 0.0, accommodating the potential overlap between true and sampled negative labels. During training, we use balanced negative sampling, ensuring that each mini-batch contains an equal number of positive and negative pairs. This approach facilitates robust model training by balancing the learning of true associations with the challenge of distinguishing potentially semantically ambiguous negatives.

#### Experiments

We evaluated the performance of CONORM across three datasets: SMM4H 2023, CADEC, and TAC. The experiments conducted for model evaluation were as follows:

1. **NER model training and evaluation**: We trained and assessed the performance of the CONORM-NER component specifically for ADE extraction on the SMM4H 2023, CADEC, and TAC datasets. For the SMM4H 2023 dataset, we employed *BERTweet* ^46^ *as the model backbone. For the CADEC dataset, we used mpnet-base* ^49^, *and for the TAC dataset, we used Clinical-Longformer* ^50^. We benchmarked the performance of CONORM-NER against state-of-the-art approaches, including MetaMap, MedCAT, GLiNER, GLiNER-bi, and JNRF.
2. **End-to-end NER and EN model training and evaluation**: We combined CONORM-NER and CONORM-EN models into an end-to-end pipeline for ADE resolution (CONORM). In the EN component, we used different pre-trained sentence transformers to generate embeddings of isolated entities and their contexts. For SMM4H 2023 and CADEC, we used *multi-qa-mpnet* ^40^ for isolated entity embeddings and *all-mpnet-base* ^40^ for context embeddings. For the TAC dataset we used *all-mpnet-base-v2* for isolated entities and *multi-qa-mpnet* for context embeddings. The performance was evaluated against MetaMap, MedCAT, and GLiNER-bi.
3. **Evaluation on undisclosed test data**: To ensure robust evaluation, we tested CONORM on undisclosed test data from the SMM4H 2023 dataset using the CodaLab platform. We compared its performance against the top-performing system from the SMM4H 2023 ADE normalization shared task ^45^, DeepADEMiner ^35^, and the average and median results of other shared task participants ^13^.

Results are reported using precision, recall, and F1-score metrics for both exact and lenient matching strategies. Performance comparisons between models were conducted using paired t-tests on per-document lenient F1-scores, with p-values *<* 0.05 considered statistically significant. This parametric approach is justified by the approximately normal distribution of score differences and the greater sensitivity of t-tests to detect mean differences when parametric assumptions are met. For each dataset, CONORM was systematically compared against the best-performing baseline in both NER and end-to-end NER and EN tasks. Statistical tests could not be performed for the undisclosed test data due to the unavailability of true labels.

## Results

The performance of CONORM was evaluated on three datasets — SMM4H 2023, CADEC, and TAC — each presenting varying degrees of language formality. CONORM consistently demonstrates strong performance across these diverse datasets, outperforming other state-of-the-art approaches in terms of F1-score for the end-to-end NER and EN task, with an average F1-score of 73.77% across the three datasets and the highest F1-score (50.20%) in the SMM4H undisclosed test set. Moreover, it achieved the highest average F1-score for both lenient and strict matching NER evaluation strategies (85.75% and 73.17%, respectively).

### Named entity recognition

Table 4 presents the NER performance of CONORM compared to state-of-the-art zero-shot (ZS) and fine-tuned (FT) systems across the SMM4H 2023, CADEC, and TAC datasets, including MetaMap, MedCAT, GLiNER-L, GLiNER-bi-L, and JNRF. On the SMM4H 2023 dataset, CONORM achieves lenient and exact F1-scores of 78.31% and 62.65%, respectively, significantly outperforming GLiNER-L (FT), which achieves F1-scores of 54.40% and 49.60% (p-value *<* 0.001). For the CADEC dataset, CONORM demonstrates its adaptability to semi-formal text with the highest lenient F1-score of 89.10% and an exact F1-score of 70.40%, showing a significant improvement over GLiNER-L (FT), which achieves 85.08% and 74.43% (p-value = 0.001). On the TAC dataset, which features formal medical documents, CONORM reaches lenient and exact F1-scores of 89.85% and 86.45%, performing competitively with GLiNER-L (FT), which achieves 90.24% and 87.99%, with no statistically significant difference observed (p-value *>* 0.05). Macro-averaged across all datasets, CONORM achieves the highest lenient and exact F1-scores among ZS and FT models, demonstrating superior generalization across diverse linguistic contexts compared to state-of-the-art methods.

**Table 4.**
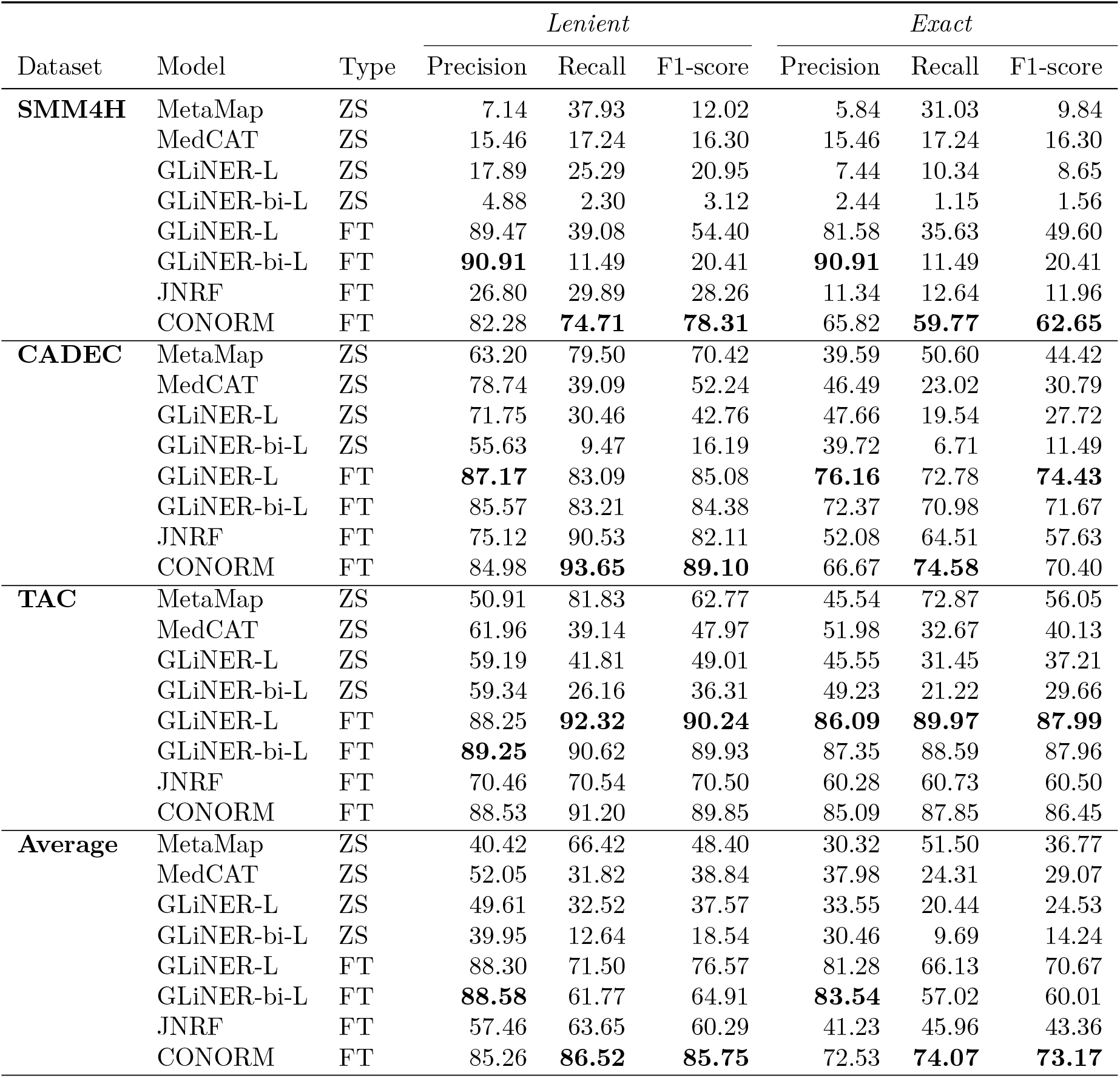
Performance comparison of CONORM and state-of-the-art systems on named entity recognition (NER) tasks across the SMM4H 2023, CADEC, and TAC datasets, evaluated using lenient and exact precision, recall, and F1-score metrics.

| Dataset | Model | Type | *Lenient* |  |  | *Exact* |  |  |
| --- | --- | --- | --- | --- | --- | --- | --- | --- |
|  |  |  | Precision | Recall | F1-score | Precision | Recall | F1-score |
| **SMM4H** | MetaMap | ZS | 7.14 | 37.93 | 12.02 | 5.84 | 31.03 | 9.84 |
|  | MedCAT | ZS | 15.46 | 17.24 | 16.30 | 15.46 | 17.24 | 16.30 |
|  | GLiNER-L | ZS | 17.89 | 25.29 | 20.95 | 7.44 | 10.34 | 8.65 |
|  | GLiNER-bi-L | ZS | 4.88 | 2.30 | 3.12 | 2.44 | 1.15 | 1.56 |
|  | GLiNER-L | FT | 89.47 | 39.08 | 54.40 | 81.58 | 35.63 | 49.60 |
|  | GLiNER-bi-L | FT | **90.91** | 11.49 | 20.41 | **90.91** | 11.49 | 20.41 |
|  | JNRF | FT | 26.80 | 29.89 | 28.26 | 11.34 | 12.64 | 11.96 |
|  | CONORM | FT | 82.28 | **74.71** | **78.31** | 65.82 | **59.77** | **62.65** |
| **CADEC** | MetaMap | ZS | 63.20 | 79.50 | 70.42 | 39.59 | 50.60 | 44.42 |
|  | MedCAT | ZS | 78.74 | 39.09 | 52.24 | 46.49 | 23.02 | 30.79 |
|  | GLiNER-L | ZS | 71.75 | 30.46 | 42.76 | 47.66 | 19.54 | 27.72 |
|  | GLiNER-bi-L | ZS | 55.63 | 9.47 | 16.19 | 39.72 | 6.71 | 11.49 |
|  | GLiNER-L | FT | **87.17** | 83.09 | 85.08 | **76.16** | 72.78 | **74.43** |
|  | GLiNER-bi-L | FT | 85.57 | 83.21 | 84.38 | 72.37 | 70.98 | 71.67 |
|  | JNRF | FT | 75.12 | 90.53 | 82.11 | 52.08 | 64.51 | 57.63 |
|  | CONORM | FT | 84.98 | **93.65** | **89.10** | 66.67 | **74.58** | 70.40 |
| **TAC** | MetaMap | ZS | 50.91 | 81.83 | 62.77 | 45.54 | 72.87 | 56.05 |
|  | MedCAT | ZS | 61.96 | 39.14 | 47.97 | 51.98 | 32.67 | 40.13 |
|  | GLiNER-L | ZS | 59.19 | 41.81 | 49.01 | 45.55 | 31.45 | 37.21 |
|  | GLiNER-bi-L | ZS | 59.34 | 26.16 | 36.31 | 49.23 | 21.22 | 29.66 |
|  | GLiNER-L | FT | 88.25 | **92.32** | **90.24** | **86.09** | **89.97** | **87.99** |
|  | GLiNER-bi-L | FT | **89.25** | 90.62 | 89.93 | 87.35 | 88.59 | 87.96 |
|  | JNRF | FT | 70.46 | 70.54 | 70.50 | 60.28 | 60.73 | 60.50 |
|  | CONORM | FT | 88.53 | 91.20 | 89.85 | 85.09 | 87.85 | 86.45 |
| **Average** | MetaMap | ZS | 40.42 | 66.42 | 48.40 | 30.32 | 51.50 | 36.77 |
|  | MedCAT | ZS | 52.05 | 31.82 | 38.84 | 37.98 | 24.31 | 29.07 |
|  | GLiNER-L | ZS | 49.61 | 32.52 | 37.57 | 33.55 | 20.44 | 24.53 |
|  | GLiNER-bi-L | ZS | 39.95 | 12.64 | 18.54 | 30.46 | 9.69 | 14.24 |
|  | GLiNER-L | FT | 88.30 | 71.50 | 76.57 | 81.28 | 66.13 | 70.67 |
|  | GLiNER-bi-L | FT | **88.58** | 61.77 | 64.91 | **83.54** | 57.02 | 60.01 |
|  | JNRF | FT | 57.46 | 63.65 | 60.29 | 41.23 | 45.96 | 43.36 |
|  | CONORM | FT | 85.26 | **86.52** | **85.75** | 72.53 | **74.07** | **73.17** |

### End-to-end entity resolution

Table 5 presents the end-to-end NER and EN performance of CONORM compared to state-of-the-art ZS and FT methods across the SMM4H 2023, CADEC, and TAC datasets. In this evaluation, we focus on computationally tractable models for large-scale entity normalization, as in the case of the MedDRA ontology. JNRF, although highly efficient, was excluded from this comparison since it is designed for NER and relation extraction rather than EN. On the SMM4H 2023 dataset, CONORM achieves an F1-score of 63.86%, significantly outperforming GLiNER-bi-L (FT), which achieves 20.63% (p-value *<* 0.001). This substantial improvement highlights CONORM’s ability to effectively process and normalize ADEs in noisy and unstructured text environments. For the CADEC dataset, CONORM attains an F1-score of 72.45%, outperforming MetaMap by 23% (p-value *<* 0.001). On the TAC dataset, consisting of formal medical documents, CONORM achieves the highest F1-score of 84.99%, with a 28% improvement over MetaMap (p-value *<* 0.001). When macro-averaged across all datasets, CONORM achieves an F1-score of 73.77%, demonstrating its superior adaptability and performance across diverse text domains compared to state-of-the-art systems.

**Table 5.** End-to-end performance comparison of CONORM and state-of-the-art systems on adverse drug event (ADE) detection and normalization tasks across the SMM4H 2023, CADEC, and TAC datasets, evaluated using precision, recall, and F1-score metrics.

| Dataset | Model | Type | Precision | Recall | F1-score |
| --- | --- | --- | --- | --- | --- |
| **SMM4H** | MetaMap | ZS | 5.19 | 27.59 | 8.74 |
|  | MedCAT | ZS | 13.40 | 14.94 | 14.13 |
|  | GLiNER-bi-L | ZS | 0.18 | 3.45 | 0.34 |
|  | GLiNER-bi-L | FT | 33.33 | 14.94 | 20.63 |
|  | CONORM | FT | **67.09** | **60.92** | **63.86** |
| **CADEC** | MetaMap | ZS | 44.17 | 56.35 | 49.53 |
|  | MedCAT | ZS | 58.35 | 28.90 | 38.65 |
|  | GLiNER-bi-L | ZS | 0.66 | 1.44 | 0.91 |
|  | GLiNER-bi-L | FT | 36.59 | 22.42 | 27.81 |
|  | CONORM | FT | **68.71** | **76.62** | **72.45** |
| **TAC** | MetaMap | ZS | 46.07 | 73.72 | 56.70 |
|  | MedCAT | ZS | 50.75 | 31.92 | 39.19 |
|  | GLiNER-bi-L | ZS | 0.19 | 0.35 | 0.25 |
|  | GLiNER-bi-L | FT | 46.12 | 38.93 | 42.22 |
|  | CONORM | FT | **83.66** | **86.37** | **84.99** |
| **Average** | MetaMap | ZS | 31.81 | 52.55 | 38.32 |
|  | MedCAT | ZS | 40.83 | 25.25 | 30.66 |
|  | GLiNER-bi-L | ZS | 0.34 | 1.75 | 0.50 |
|  | GLiNER-bi-L | FT | 38.68 | 25.43 | 30.22 |
|  | CONORM | FT | **73.15** | **74.64** | **73.77** |

### End-to-end entity resolution: undisclosed test data

To further validate the effectiveness of CONORM in real-world settings, we evaluated our model on the undisclosed test set of the SMM4H 2023 dataset using the CodaLab platform. Table 6 presents the comparative performance of CONORM against other leading systems, including the previous top-performing system from the SMM4H 2023 ADE normalization shared task ^45^ and DeepADEMiner ^35^. The table details both the overall and the out-of-distribution performance metrics. CONORM achieved an overall F1-score of 50.20%, which outperforms both the previous top system and DeepADEMiner. Importantly, CONORM achieves the highest precision (53.50%). This means that our model can identify ADEs with fewer false positives as compared to top-performing models. Although DeepADEMiner achieved a slightly higher recall, CONORM’s balanced precision and recall led to a superior F1-score (5% improvement). In terms of out-of-distribution performance, which evaluates the model’s ability to generalize to ADEs not seen during training, CONORM again outperforms other systems with an F1-score of 39.40% and a precision of 53.10%. These results demonstrate that CONORM achieves state-of-the-art performance for ADE resolution in informal tweet data, particularly in terms of precision and generalization to out-of-distribution cases.

**Table 6.** Performance comparison of CONORM, DeepADEMiner^35^, the top-performing system from the SMM4H 2023 ADE normalization shared task 5^45^, and the mean and median results of shared task participants ^13^ on the SMM4H 2023 undisclosed test set. Metrics include overall and out-of-distribution (OOD) precision, recall, and F1-score.

| Model | *Overall performance* |  |  | *OOD performance* |  |  |
| --- | --- | --- | --- | --- | --- | --- |
|  | Precision | Recall | F1-score | Precision | Recall | F1-score |
| Mean of participants | 29.30 | 42.24 | 32.94 | 15.08 | 36.02 | 20.16 |
| Median of participants | 24.90 | 40.50 | 32.20 | 12.80 | 35.40 | 19.50 |
| Top system<sup>45</sup> | 44.90 | 40.50 | 42.60 | 24.90 | 35.40 | 29.20 |
| DeepADEMiner<sup>35</sup> | 40.70 | **50.80** | 45.20 | 33.50 | **39.50** | 36.30 |
| **CONORM (ours)** | **53.50** | 47.30 | **50.20** | **53.10** | 31.40 | **39.40** |

## Model analysis and characterization

### Adaptive balancing of isolated and contextual information

We conducted a study to assess how CONORM-EN balances isolated and contextual embeddings across datasets with varying linguistic formality. DCR learns a weighting function during training, which dynamically generates *λ* (for isolated embeddings) and *µ* (for context embeddings) at inference time, adapting these weights to each input instance.

To evaluate how well DCR generalizes, we compared the average *µ* values from the DCR mechanism to an oracle using fixed *µ* values. In this setup, we combined pre-trained isolated and context embeddings using static weights ranging from 0 to 1, applied post-hoc on the test set, representing upper-bound performance estimates.

Figure 2A presents end-to-end F1-scores across fixed *µ* values, along with DCR’s *µ* averages. As shown in Figure 2A, DCR consistently learns near-optimal values. For TAC, the average *µ* = 0.38 aligns with the optimal *µ ∈* [0.25, 0.50], indicating the importance of isolated embeddings for formal entity mentions in medical texts. For CADEC, DCR generates an average *µ* of 0.88, close to the oracle peak around *µ≈* 0.75, strongly relying on contextual cues with support from isolated representations. In SMM4H, the most informal dataset, DCR learned to leverage context with a high *µ* = 0.82, closely matching the oracle (*µ* = 1.0), which suggests that context is key for entity disambiguation.

**Figure 2.**
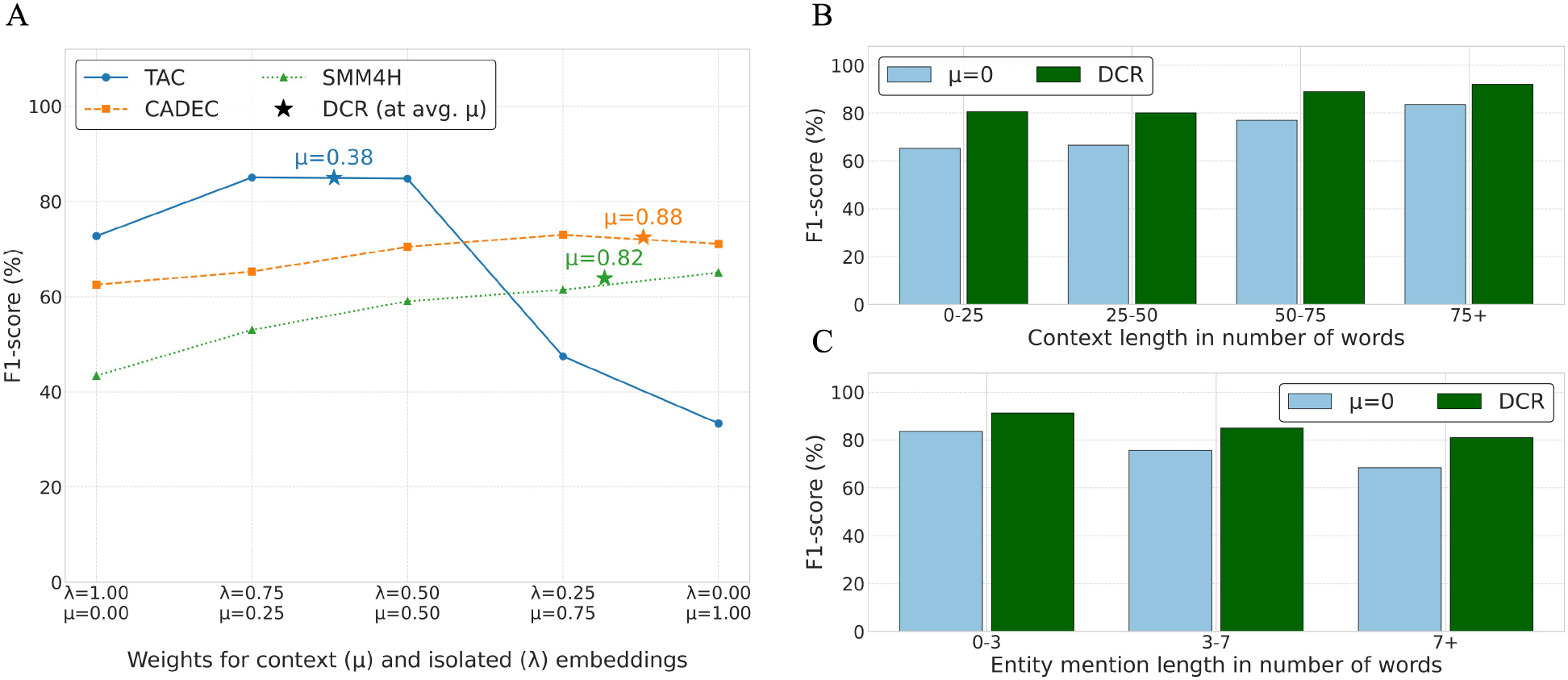
Analysis of CONORM’s end-to-end entity recognition and normalization performance using F1-score. (A) Performance sensitivity to static weighting between context (*µ*) and isolated (*λ*) embeddings across TAC, CADEC, and SMM4H test sets. Lines track F1-score as *µ* varies from 0.0 (isolated-only) to 1.0 (context-only). Stars indicate the performance using the learned DCR, plotted at the dataset-specific average *µ* computed at inference time. (B) Performance on the TAC test set for isolated-only (*µ* = 0) and DCR configurations, stratified by the context length containing the target entity to normalize. (C) Conditional F1-score for the same configurations and dataset as (B), but stratified by the extracted entity mention length. This score assesses CONORM-EN’s performance given a mention of a certain length was extracted by CONORM-NER.

This analysis reveals two key findings. First, DCR learns to approximate near-optimal weighting solely from training data, without relying on test-time tuning. Second, it adapts the contribution of each encoder based on text formality. In formal domains, it leans on isolated embeddings to exploit pre-trained knowledge. In informal settings, it emphasizes learned contextual cues while still leveraging general-domain semantics. This dynamic context refining is central to CONORM-EN’s robust performance across different text formalities.

### Performance across varying context and mention lengths

The subsequent in-depth analyses exclusively utilize the TAC dataset. This dataset, featuring formal language within long documents, was chosen for its large volume of annotations and balanced distribution across clinical categories, making it ideal for stratified performance evaluations of factors like context utilization, out-of-distribution generalization, and performance variations across system organ classes.

To further understand CONORM-EN’s behaviour, we analyzed its performance on the TAC test set when stratified by context length (Figure 2B) and extracted entity mention length (Figure 2C). We compare the DCR mechanism against the isolated-only (*µ* = 0) embedding strategy, which represents end-to-end model normalization performance in the absence of contextual cues.

As shown in Figure 2B, the isolated-only configuration shows improved performance with longer contexts, perhaps due to better entity boundary detection by CONORM-NER in longer contexts. Notably, the DCR configuration consistently outperforms the context-free baseline across all context lengths. It demonstrates an important performance increase in longer passages, showcasing its ability to effectively utilize extended context when available, and synergize it with isolated information better than the isolated-only configuration.

Figure 2C examines the conditional F1-score based on the length of the extracted entity itself. This metric assesses normalization accuracy given that CONORM-NER identified a mention. A general trend observed across all configurations is a decrease in performance as mention length increases, suggesting that longer multi-word expressions are inherently more challenging to normalize. However, the differences between DCR and *µ* = 0 are stark. Isolated-only (*µ* = 0) performs relatively well, highlighting the importance of isolated embeddings’ general knowledge for normalizing mentions regardless of length. CONORM-EN with DCR achieves the highest conditional F1-score across all mention lengths. This suggests DCR effectively adapts its weighting strategy not only based on broader context but also in response to the characteristics of the mention itself, leading to more robust normalization across varying mention complexities.

### Generalization to out-of-distribution concepts

To assess CONORM-EN’s ability to generalize to concepts unseen during training, we stratified the evaluation of the TAC test set according to whether the gold-standard MedDRA concept for each entity appeared in the training data. We compared CONORM-EN using the DCR mechanism against the isolated-only (*µ* = 0) baseline. The resulting precision and recall values, along with F1-scores, are presented in Figure 3.

**Figure 3.**
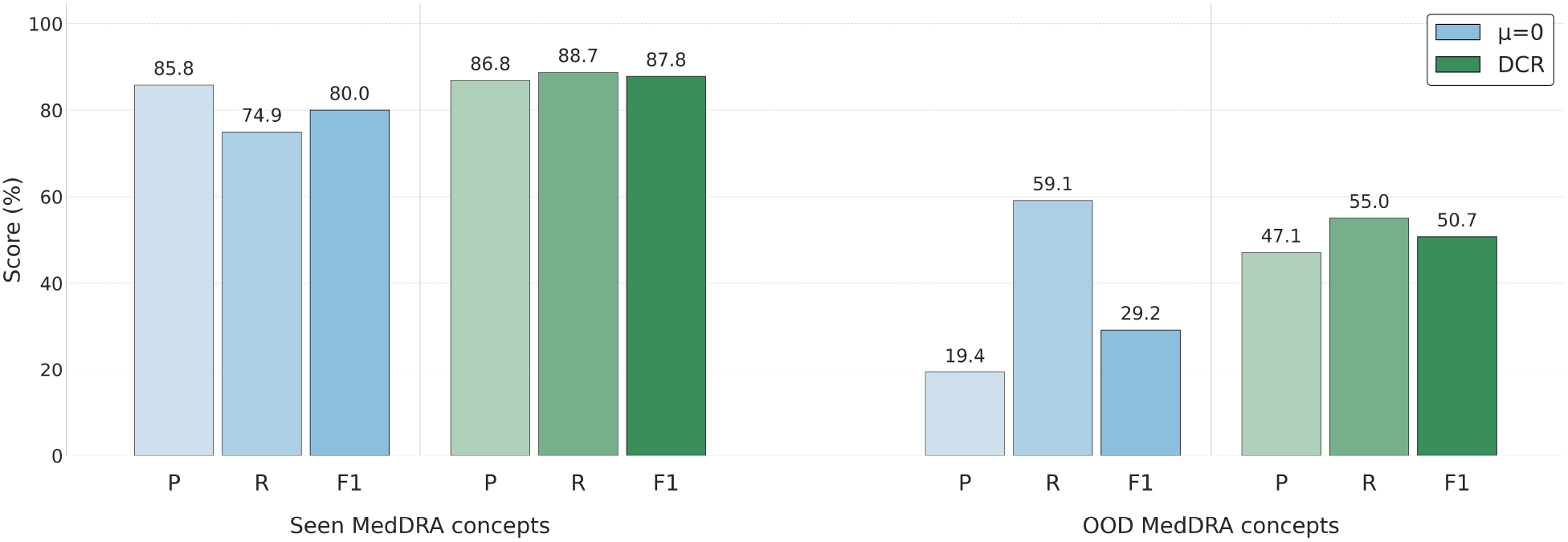
Analysis of end-to-end performance on the TAC test set, stratified by MedDRA concept exposure during training. The figure compares precision (P), recall (R), and F1-score (F1) for CONORM under isolated-only (*µ* = 0) and DCR configurations, evaluated separately on seen and out-of-distribution (OOD) MedDRA concepts.

As illustrated in Figure 3, both configurations perform substantially better on seen MedDRA concepts compared to out-of-distribution (OOD) concepts, highlighting the inherent difficulty in normalizing novel medical terms. For concepts previously encountered during training (seen concepts), the isolated-only configuration (*µ* = 0) maintains relatively high precision (85.8%), but its recall (74.9%) is noticeably lower than that achieved by DCR. Incorporating DCR substantially improves recall, demonstrating DCR’s effectiveness at leveraging contextual information for previously encountered concepts.

Performance differences become more pronounced when examining OOD concepts. The isolated-only configuration (*µ* = 0) exhibits relatively high recall (59.1%), reflecting the ability of isolated embeddings to capture general semantic information learned during transformer pre-training. However, its precision remains low (19.4%), indicating frequent incorrect normalizations of novel terms. Employing DCR notably improves precision (47.1%), achieving a gain of +27.7 percentage points compared to the isolated-only configuration, while slightly reducing recall (55.0%). Thus, on OOD terms, DCR’s primary contribution shifts from enhancing recall, as observed with in-distribution terms, to substantially improving precision by effectively disambiguating novel medical concepts.

In summary, normalizing OOD concepts remains inherently challenging, but CONORM-EN employing the DCR mechanism demonstrates meaningful improvements in generalization. By dynamically adjusting the influence of context, DCR proves versatile: it effectively enhances recall for seen concepts and substantially improves precision for unseen concepts. This highlights its value in robustly normalizing novel medical terminology encountered post-training.

### Prediction flips

We investigate the specific impact of the DCR mechanism in CONORM-EN by analyzing how its predictions change relative to deactivated context (*µ* = 0) on the TAC test set (Table 7). This analysis reveals how incorporating dynamic context influences the baseline’s normalization outcome for correctly or incorrectly normalized entities.

**Table 7.** Analysis of prediction flips from CONORM-EN with *µ* = 0 to CONORM-EN with DCR on the TAC test set. Percentages are relative to the baseline model’s predictions.

| Category | Count | Percentage |
| --- | --- | --- |
| *True positives (baseline, $\mu = 0$)* |  |  |
| Total | 8245 | - |
| $\hookrightarrow$ Kept correct (TP → TP) | 8149 | 98.84% |
| $\hookrightarrow$ Became missed (TP → FN) | 96 | 1.16% |
| *False negatives (baseline, $\mu = 0$)* |  |  |
| Total | 2929 | - |
| $\hookrightarrow$ Corrected by DCR (FN → TP) | 1491 | 50.90% |
| $\hookrightarrow$ Remained missed (FN → FN) | 1438 | 49.10% |

Examining the 8245 true positives (TPs) identified when *µ* = 0, we observe high stability when DCR is applied. Among all baseline TPs remain correctly normalized (98.84%). This indicates that the introduction of context rarely disrupts an already correct prediction based on isolated information alone. Conversely, focusing on the 2929 false negatives (FNs) produced when *µ* = 0, DCR demonstrates a substantial corrective capability. It successfully flips 50.90% of these baseline errors into TPs. This clearly illustrates the benefit of dynamically incorporating contextual embeddings to resolve cases where isolated information is insufficient or ambiguous.

Overall, this analysis shows that CONORM-EN’s DCR successfully corrects over half of the FNs made when relying solely on isolated embeddings (50.90%) and introduces a small number of new errors (1.16%). The substantial net gain underscores the effectiveness of CONORM-EN’s dynamic weighting approach.

### Performance across SOCs

Assessing the performance of ADE detection and normalization models across different medical categories is crucial for understanding their practical utility in pharmacovigilance. MedDRA’s SOCs encompass diverse medical conditions, each with unique linguistic features and terminological complexities. To explore these factors, we conducted a detailed analysis of CONORM’s end-to-end performance using the TAC dataset, stratifying the results by SOC in the test set.

As presented in Table 8, CONORM’s performance varies across the 25 different SOCs present in the test data. It achieves an F1-score equal to or above 80% on 19 out of these 25 categories. Notably high performance is observed for *Gastrointestinal* (94.95%) and *Ear and labyrinth* (94.87%) disorders. In contrast, lower F1-scores are seen in SOCs with lower support, such as *Social circumstances* (44.44%) and *Congenital* disorders (76.92%). Furthermore, a Spearman rank correlation analysis revealed a statistically significant positive association between the training support for an SOC and its F1-score (*ρ* = 0.5, p-value *<* 0.05), suggesting that categories with more representation in the training data tend to exhibit better performance.

**Table 8.**
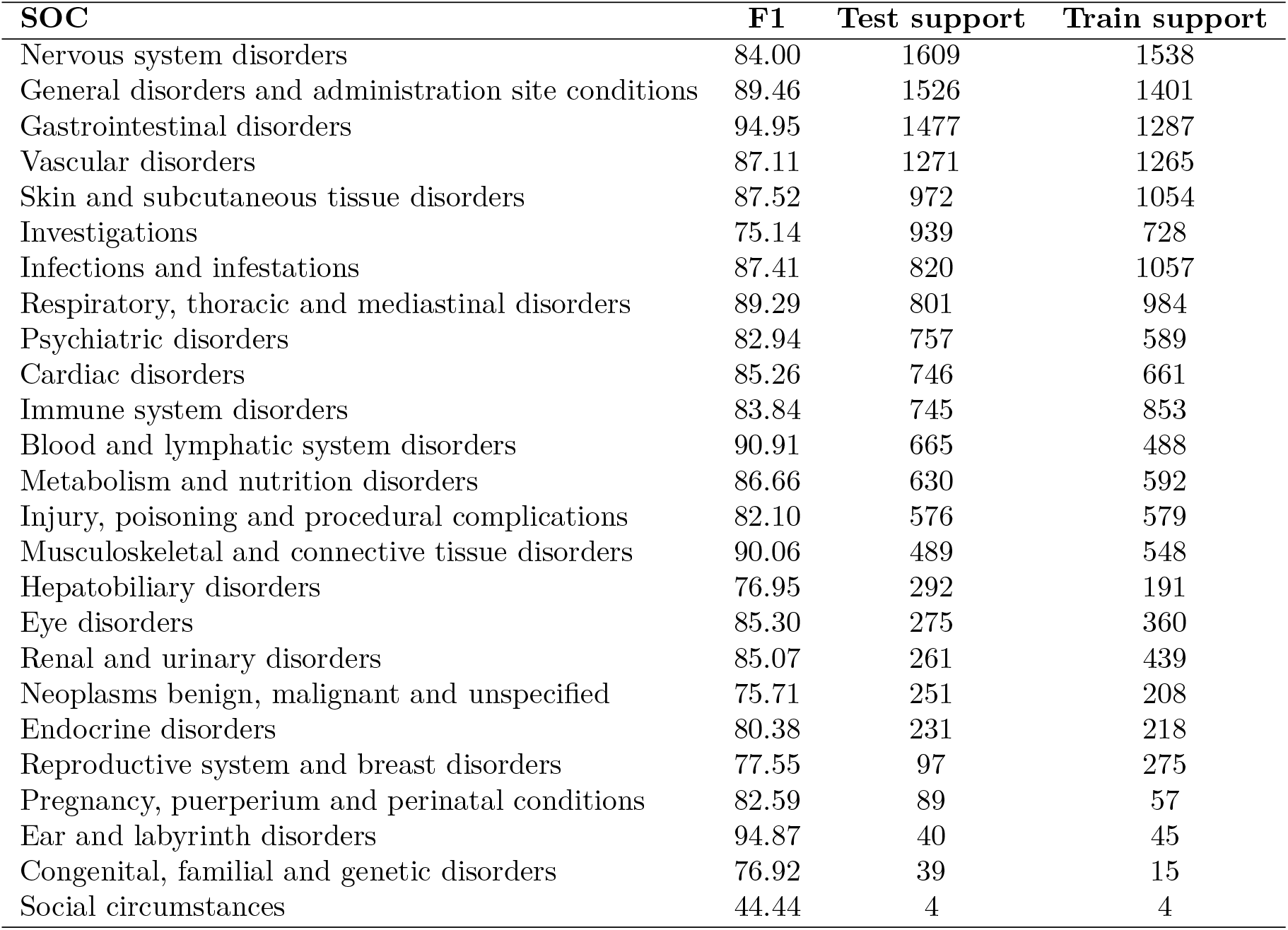
Stratified performance of CONORM on end-to-end entity resolution for the TAC dataset, evaluated by system organ class (SOC). F1-scores and support counts for test and train sets are shown.

| SOC | F1 | Test support | Train support |
| --- | --- | --- | --- |
| Nervous system disorders | 84.00 | 1609 | 1538 |
| General disorders and administration site conditions | 89.46 | 1526 | 1401 |
| Gastrointestinal disorders | 94.95 | 1477 | 1287 |
| Vascular disorders | 87.11 | 1271 | 1265 |
| Skin and subcutaneous tissue disorders | 87.52 | 972 | 1054 |
| Investigations | 75.14 | 939 | 728 |
| Infections and infestations | 87.41 | 820 | 1057 |
| Respiratory, thoracic and mediastinal disorders | 89.29 | 801 | 984 |
| Psychiatric disorders | 82.94 | 757 | 589 |
| Cardiac disorders | 85.26 | 746 | 661 |
| Immune system disorders | 83.84 | 745 | 853 |
| Blood and lymphatic system disorders | 90.91 | 665 | 488 |
| Metabolism and nutrition disorders | 86.66 | 630 | 592 |
| Injury, poisoning and procedural complications | 82.10 | 576 | 579 |
| Musculoskeletal and connective tissue disorders | 90.06 | 489 | 548 |
| Hepatobiliary disorders | 76.95 | 292 | 191 |
| Eye disorders | 85.30 | 275 | 360 |
| Renal and urinary disorders | 85.07 | 261 | 439 |
| Neoplasms benign, malignant and unspecified | 75.71 | 251 | 208 |
| Endocrine disorders | 80.38 | 231 | 218 |
| Reproductive system and breast disorders | 77.55 | 97 | 275 |
| Pregnancy, puerperium and perinatal conditions | 82.59 | 89 | 57 |
| Ear and labyrinth disorders | 94.87 | 40 | 45 |
| Congenital, familial and genetic disorders | 76.92 | 39 | 15 |
| Social circumstances | 44.44 | 4 | 4 |

## Discussion

We introduced CONORM, a context-aware entity normalization framework specifically designed for the detection and normalization of ADEs from unstructured text sources. CONORM addresses key challenges in processing informal, semi-formal, and formal text domains. The effectiveness of the DCR mechanism, a core component of CONORM-EN, is detailed in our analyses (Section), demonstrating its ability to adaptively balance information sources and significantly contribute to performance across diverse text domains. Our comprehensive evaluation across three benchmark datasets - SMM4H 2023, CADEC, and TAC - demonstrated that CONORM consistently achieves superior or competitive performance compared to state-of-the-art methods in both NER and end-to-end ADE recognition and normalization tasks. These results position CONORM as a robust and versatile solution for ADE resolution, capable of adapting to diverse textual domains and linguistic complexities.

In the NER task, CONORM exhibited strong performance across all datasets. The difference between lenient and exact F1-scores provides insights into the model’s precision in boundary detection. On the informal SMM4H dataset, CONORM achieved a lenient F1-score of 78.31% but a lower exact F1-score of 62.65%. This gap highlights the challenges in identifying ADE boundaries in texts with slang, abbreviations, and misspellings. In contrast, the formal TAC dataset showed a smaller difference between lenient (89.85%) and exact (86.45%) F1-scores, reflecting the relatively simpler boundary detection in structured medical documents. Compared to rule-based models like MetaMap, CONORM’s transformer-based architecture captures contextual information, enabling better generalization to diverse expressions of ADEs. Rule-based systems rely on predefined vocabularies and patterns, making them less adaptable to non-standard expressions and contextual cues.

When comparing our NER results to related work, CONORM demonstrates competitive performance. On the CADEC dataset, CONORM achieved an exact F1-score of 70.40%, slightly lower than the 72.99% reported by Ul Haq *et al*. ^41^, who used micro-averaged metrics across both drug and ADE entities. However, CONORM focuses solely on continuous ADE mentions, making a direct comparison challenging. CONORM outperformed in lenient F1-score (89.10%) compared to Ul Haq *et al*.’s 86.88%, indicating its strength in maintaining high recall and precision, particularly in semi-formal texts. For the TAC dataset, CONORM achieved an exact F1-score of 86.45%, comparable to the 85.20% reported by UTH CCB ^17^, which focused on all ADE mentions. Although UTH CCB did not report lenient metrics, CONORM’s lenient F1-score (89.85%) underscores its effectiveness in formal text environments.

In the end-to-end entity resolution task, CONORM demonstrated its efficacy in both extracting ADE mentions and accurately normalizing them to MedDRA terms. The model consistently outperformed other systems across all datasets, with particular improvements on the SMM4H 2023 dataset. An F1-score as high as 63.86% on the informal SMM4H dataset indicates CONORM’s robustness in challenging linguistic environments, supported by DCR’s demonstrated ability to adapt its weighting strategy based on text formality, emphasizing contextual information more heavily in such informal settings (Section). On the undisclosed test data from SMM4H 2023, CONORM also achieved the highest overall and out-of-distribution F1-scores (50.20% and 39.40%, respectively), outperforming previous top systems (Table 6). This strong performance, particularly the high precision observed even on out-of-distribution concepts (Section, Section), highlights the benefit of DCR in reducing false positives, which is critical for clinical applications where over-reporting may result in unnecessary interventions.

A key component contributing to CONORM’s performance is the dynamic context refining (DCR) mechanism within CONORM-EN. While leveraging context and attention concepts is established in NLP, DCR’s specific contribution lies in its learned, dynamic weighting within a dual-encoder normalization framework. Our analyses underscore the practical value of this approach. DCR learns near-optimal weighting based on text formality (Section), substantially corrects errors made by context-agnostic approaches by successfully resolving over half of their false negatives (Section), and crucially improves precision when normalizing novel, out-of-distribution medical concepts (Section). This adaptive integration of pre-trained semantic knowledge with task-specific contextual cues, informed by the surrounding text and the mention itself, appears central to CONORM’s robust cross-domain performance.

Comparing our end-to-end results to related work further highlights CONORM’s effectiveness. Remy *et al*. ^44^ focused solely on entity normalization and reported an F1-score of 60.28% on the CADEC dataset using BioLORD-STS. In contrast, CONORM addresses both NER and EN in an end-to-end pipeline and achieves an F1-score of 72.45%. On the TAC dataset, UTH CCB ^17^ achieved a precision of 84.17%, recall of 89.84%, and F1-score of 86.91% by combining NER and EN; however, their evaluation process only required one correct detection and normalization per unique entity per document to count as a true positive, regardless of how many times that entity appeared. In contrast, we evaluated every occurrence of each entity, making our evaluation stricter. Under this criterion, CONORM achieved a precision of 83.66%, recall of 86.37%, and F1-score of 84.99%.

Our detailed model analysis (Section) provides further insights into CONORM’s performance dynamics and highlights important considerations for ADE detection and normalization. The variation in performance across SOCs (Table 8) may be attributed to differences in the complexity of medical terminology, condition specificity, and the linguistic expressions used to describe these categories. CONORM achieves notably high performance for SOCs such as *Gastrointestinal* and *Blood and lymphatic* disorders, suggesting that these more frequently discussed ADEs are easier to detect and normalize. Conversely, performance is lower in less common SOCs such as *Social circumstances*. Our analysis confirmed a statistically significant positive correlation between training support and F1-score per SOC (Spearman’s *ρ* = 0.5, p-value *<* 0.05), highlighting the impact of data availability. Examining performance across varying input characteristics (Section) further illuminates the role of context and mention structure. Our analysis shows that CONORM with DCR effectively utilizes context, demonstrating robustness in longer passages where simpler approaches might underperform (Figure 2B). While performance tends to decrease for longer entity mentions, DCR consistently outperforms the context-free baseline, indicating its effective integration of information regardless of mention complexity (Figure 2C).

Although CONORM has demonstrated high performance across informal, semi-formal, and formal text domains, there are limitations to this work that warrant consideration. First, the SMM4H dataset had relatively few ADE annotations, restricting the diversity of examples available for model training. Nonetheless, the aggregated number of samples across SMM4H, CADEC, and TAC remains robust, supporting a comprehensive evaluation. Second, each model was tailored to its respective dataset, yet a single unified model that simultaneously learns from all three datasets might further improve generalization. We leave this exploration for future research. Finally, as all datasets are in English, any extension of CONORM to other languages would require additional adaptation and training, underscoring an avenue for broader multilingual applications.

Future work could address these limitations in several ways. First, developing or collecting augmented and synthetic data may alleviate the scarcity of instances in underrepresented SOCs, improving resolution performance, particularly for categories identified as challenging in our analysis (Section). Second, extending CONORM to handle multilingual corpora would enhance the framework’s relevance for global pharmacovigilance. Additionally, deploying CONORM on streaming social media data could enable near real-time monitoring, making it useful for rapid ADE detection during public health crises or newly released drugs. Finally, improving computational efficiency, such as through model quantization, could facilitate low-latency inference in resource-constrained environments.

## Conclusion

In this work, we introduced CONORM, an end-to-end framework designed to improve adverse drug event (ADE) detection and normalization across diverse text sources, from informal social media to formal product labels. A key contribution is the dynamic context refining (DCR) mechanism within CONORM’s dual-encoder normalization module. Our comprehensive analyses provided strong empirical evidence for DCR’s effectiveness. DCR adaptively balances isolated entity semantics and contextual information based on text formality, enhances precision for challenging out-of-distribution concepts, and substantially corrects errors made by context-agnostic baseline models. Leveraging transformer-based models and DCR, CONORM consistently achieved state-of-the-art performance across benchmark datasets, demonstrating robust recognition of ADE boundaries and accurate normalization to MedDRA terms across varying linguistic styles. The framework’s scalable inference methodology further ensures its practicality for large-scale pharmacovigilance applications involving extensive medical terminologies. While our experiments highlight strong general performance and adaptability, future research could refine the framework for multilingual scenarios and further address challenges posed by scarce system organ classes. By delivering improved accuracy and robustness in ADE resolution, particularly through its context-aware normalization strategy, CONORM can help advance drug safety monitoring, informing public health strategies and clinical decision-making processes aimed at reducing medication-related harm.

## Data Availability

The datasets supporting the results of this article are available as follows: The CADEC dataset can be accessed at https://data.csiro.au/collection/csiro:10948. The TAC dataset is available at https://bionlp.nlm.nih.gov/tac2017adversereactions/. The SMM4H 2023 dataset is available upon request at https://codalab.lisn.upsaclay.fr/competitions/12941#participate-get-data. All datasets are provided in their raw formats, and preprocessing scripts to replicate our experiments and prepare the data for the CONORM framework are available at https://github.com/ds4dh/CONORM. The MedDRA files used for entity normalization and data preprocessing can be obtained upon request from https://www.meddra.org/. Specifically, we used MedDRA English version 16.0 for CADEC, version 24.0 for SMM4H 2023, and version 18.1 for TAC, ensuring consistency with the original dataset annotations.

https://bionlp.nlm.nih.gov/tac2017adversereactions/

https://codalab.lisn.upsaclay.fr/competitions/12941#participate-get-data

https://www.meddra.org/

## Code and data availability

The datasets supporting the results of this article are available as follows: The CADEC ^16^ dataset can be accessed at CADEC. The TAC ^17^ dataset is available at TAC. The SMM4H 2023 dataset ^13^ is available upon request at SMM4H 2023. All datasets are provided in their raw formats, and preprocessing scripts to replicate our experiments and prepare the data for the CONORM framework are available at https://github.com/ds4dh/CONORM. The MedDRA files used for entity normalization and data preprocessing can be obtained upon request from https://www.meddra.org/. Specifically, we used MedDRA English version 16.0 for CADEC, version 24.0 for SMM4H 2023, and version 18.1 for TAC, ensuring consistency with the original dataset annotations.

## Author contributions

AY and DT contributed to the study’s conceptualization. AY, HR, and AB developed the methodology. AY carried out the experiments and wrote the original draft. AB, HR, and DT reviewed and edited the manuscript, with supervision provided by DT.

## Acknowledgment

This work was funded by the Innosuisse - project no.: 55441.1 IP ICT.

